# Community-level women’s education and hemoglobin level among Indian adolescents: A multilevel analysis of a national survey

**DOI:** 10.1101/2020.11.04.20224154

**Authors:** Ankita R. Shah, Malavika A. Subramanyam

## Abstract

**Background:** Little research has explored the influence of social context on hemoglobin levels among Indian adolescents. We conceptualized community-level women’s education (proxy for value placed on women’s wellbeing) as exerting contextual influence on adolescent hemoglobin level.

**Methods:** We used the Indian National Family Health Survey 2015-16 data (62,648 adolescents aged 15-17 years). We fit multilevel random intercepts linear regression models to test the association of village- and urban-ward-level-women’s education with hemoglobin of adolescents, accounting for own and their mother’s education; plus relevant covariates.

**Findings:** Our fully adjusted model estimated that if the 45% of communities with fewer than ten percent of women having a tenth-grade education in our sample were to achieve 100% high school completion in women, the average hemoglobin of all adolescents in such communities would be 0·22 g/dl higher (p<0·01). Unexplained variance at the contextual level remained statistically significant, indicating the importance of context on adolescent hemoglobin.

**Interpretations:** Adolescents are deeply embedded in their context, influenced by contextual factors affecting health. Promoting adolescent health therefore implies altering social norms related to adolescent health and health behaviors; along with structural changes creating a health-promoting environment. Integrating our empirical findings with theoretically plausible understanding of the pathways connecting community-level women’s education with adolescent hemoglobin, we suggest that enhancing community-level women’s education beyond high school is necessary to facilitate these processes.

**Implications:** Addressing contextual determinants of adolescent hemoglobin might be the missing link in India’s adolescent anemia prevention efforts, which are focused heavily on individual-level biomedical determinants of the problem.

## Introduction

Low levels of hemoglobin and anemia are pressing health risks in adolescents (10 to 19 years) and demand urgent action.^1^ With 253 million adolescents, roughly a quarter of its population, India is home to the world’s largest number of adolescents.^2^ Anemia among Indian adolescents affects 40% of girls and 18% of boys.^2^ Anemia severely curbs growth and cognitive development, adversely affects productivity, and increases the risk of poor pregnancy outcomes among adolescent mothers.^2^

Despite such severe impacts, the research on low hemoglobin levels and anemia in Indian adolescents is limited in the theoretical lens employed. While a range of individual-level social, behavioral, biological and physiological risk factors associated with anemia in adolescent girls are reported,^34^ there is a paucity of research examining the potential impact of context on anemia in adolescents.

Socio-economic and political context, including cultural and societal values and social stratification based on gender, caste, class and religion, are important social determinants of nutrition in adolescents.^5^ They operate at global, regional, and local levels through complex intersecting pathways, determining access to and availability of food, health, education and security to adolescents living in nested social hierarchies.^5^ An individual-focused perspective fails to take into account the causal influence of the context on adolescents’ hemoglobin levels.^6^ We conceptualize that living in neighborhoods that support sociocultural norms valuing women’s well-being promote adolescent health. We investigate this by quantifying the association of community-level women’s education with adolescent hemoglobin level, above and beyond the role of their mother’s education.

To our knowledge, this is the first study that investigates the relationship of community-level women’s education with adolescents’ nutritional status.

## Methods

### Dataset and sample characteristics

The analysis is based on publicly available, de-identified data from the 2015-16 round of the National Family Health Survey (NFHS-4), India, undertaken in 2015-16.^7^ It was a nationally representative survey, giving estimates at district and state levels.^8^ The sampling design treated villages (occasionally village-clusters) and census-enumeration wards (in urban areas) as the Primary Sampling Units (PSUs),^8^ which we operationalize as the “community.”

A total of 601,509 households were interviewed, which were selected by two-stage stratified random sampling,^8^ (for additional details on sampling design refer^8^) from 640 districts of all 36 states/union territories (UT) in India were included in the survey. Hemoglobin was measured in a total of 117,711 girls and 17,912 boys aged 15 through 19 years. We restricted our analysis to unmarried adolescents between 15 through 17 years of age, as information on parents’ education was available only for the children below 18 years of age.^8^ We linked hemoglobin levels of 75,696 adolescents to their mother’s education level. As information on father’s education was collected only if the father permanently lived in the same household as that of the adolescents^8^ (information not available for instance if father had migrated for work), we excluded 11,106 adolescents who did not have the information on father’s education. Further excluding observations having missing data (including the response “don’t know”) on any of the other covariates, our analytical sample size was 62,648 adolescents.

### Measures

Our main predictor was education of women at the community-level, represented as the proportion of all women in NFHS-4 above 19 years of age in a PSU who had completed formal education at least till the tenth grade. Education data of a total of 946,297 women across PSUs were used to compute this variable. The outcome variable was blood hemoglobin level in adolescents measured in grams per deciliter (g/dl), on a continuous ratio scale. Hemoglobin estimation was conducted on-site with a battery-operated portable HemoCue Hb 201+ analyser.^8^ Hemoglobin levels were adjusted for smoking and altitude in areas that were above 1,000 meters.^8^

Data on age (years), gender (girls or boys), religion (Hindu, Muslim, Christian, and others), social group (scheduled tribe-ST, scheduled caste-SC, other backward classes-OBC, and general), household wealth (quintiles based on the score on the first principal component of a household asset score), place of residence (urban or rural), household size, education of adolescents, their mother and father (all as years of formal schooling completed), and sub-national regions (groups of states in the North, East, West, Central, South and Northeast) were also included.^8^ Terminology used to denote social groups is adopted by the Government of India, where ST, SC and OBC represent historically underprivileged groups. Variables representing the socio-economic condition of the community were created as the proportions, respectively, of total households in the PSUs following Muslim religion, belonging to ST/SC groups and the poorest wealth category.

### Statistical analysis

Descriptive statistics were employed to depict the distribution of the sample, and hemoglobin levels, across the covariates. We fitted five-level random intercept multiple linear regression models to estimate the association of community-level women’s education with adolescent hemoglobin levels. This was an appropriate choice given our interest in the influence of the context on an individual-level outcome.^9^ The five hierarchical population levels were adolescent *i*, nested in household *j*, PSU (community) *k*, district *l* and state *m*. The random components at each level were assumed to follow a normal distribution, with a mean of zero, and a constant variance; and assumed to be mutually independent. Alpha was set at 0·05. The unadjusted Model 1 included community-level women’s education as the predictor. We incrementally added individual-level covariates: adolescent’s gender and age in Model 2, adolescent’s education in Model 3, and mother’s education in Model 4. Fully adjusted Model 5 additionally included father’s education, household-level covariates: family size, social group, religion, household wealth; community-level covariates: wealth, and social and religious composition; and state-level covariate: region. Statistical analysis was carried out using STATA, version 12·1.^10^

## Results

Our analytical sample consisted of a total of 62,648 adolescents aged 15 to 17 years (average: 16·45; SD: 0·89), from 57,638 households, 23,448 PSUs, 640 districts, and 36 states/UT. The sample consisted mainly of girls (87%) (Table 1). Distribution of adolescents by background characteristics in our sample was comparable with that of the Indian adolescent population (Census 2011);^11^ except for the social group. In contrast to 9·2% of the adolescents belonging to ST group in India, 17·5% adolescents in our sample belonged to the ST group; whereas the adolescents in the general category were lower than the national average.^11^ Community-level descriptive statistics are given in table 2.

**Table 1:** Distribution of the adolescents and average hemoglobin levels (N= 62,648) by gender and sociodemographic characteristics

|  | Girls |  | Boys |  | Total<br>n (column %) |
| --- | --- | --- | --- | --- | --- |
|  | n (column %) | Hemoglobin<br>level<br>Mean (SD) | n (column %) | Hemoglobin<br>level<br>Mean (SD) |  |
| Religion |  |  |  |  |  |
| Hindu | 41255 (76.07) | 11.68 (1.56) | 6482 (77.02) | 13.47 (1.68) | 47737 (76.20) |
| Muslim | 7334 (13.52) | 11.74 (1.57) | 1069 (12.70) | 13.56 (1.69) | 8403 (13.41) |
| Christian | 3454 (6.37) | 12.32(1.56) | 482 (5.73) | 14.09 (1.69) | 3936 (6.28) |
| Other | 2189 (4.04) | 11.66 (1.65) | 383 (4.55) | 13.52 (1.65) | 2572 (4.11) |
| Caste background |  |  |  |  |  |
| Scheduled Caste | 10966 (20.22) | 11.59 (1.61) | 1705 (20.26) | 13.43 (1.74) | 12671 (20.23) |
| Scheduled Tribe | 9549 (17.61) | 11.75 (1.62) | 1419 (16.86) | 13.48 (1.72) | 10968 (17.51) |
| other backward<br>caste | 23076 (42.55) | 11.72 (1.54) | 3525 (41.88) | 1.52 (1.67) | 26601 (42.46) |
| General | 10641 (19.62) | 11.85 (1.54) | 1767 (21.00) | 13.64 (1.64) | 12408 (19.81) |
| Wealth quintile |  |  |  |  |  |
| Poorest | 12126 (22.36) | 11.57 (1.56) | 1577 (18.74) | 13.17 (1.64) | 13703 (21.87) |
| Poorer | 13068 (24.10) | 11.72 (1.58) | 1963 (23.32) | 13.41 (1.65) | 15031 (23.99) |
| Middle | 11550 (21.30) | 11.77 (1.57) | 1787 (21.23) | 13.57 (1.72) | 13337 (21.29) |
| Richer | 9624 (18.00) | 11.76 (1.57) | 1655 (19.66) | 13.65 (1.73) | 11279(18.00) |
| Richest | 7864 (14.84) | 11.86 (1.55) | 1434 (17.04) | 13.83 (1.65) | 9298 (14.84) |
| Place of residence |  |  |  |  |  |
| urban | 14479 (26.70) | 11.80 (1.57) | 2437 (28.96) | 13.71 (1.71) | 16916 (27.00) |
| rural | 39753 (73.30) | 11.70 (1.57) | 5979 (71.04) | 13.44 (1.68) | 45 |
|  |  |  |  |  | 732 (73.00) |
| Region |  |  |  |  |  |
| North | 16627 (30.66) | 11.66 (1.61) | 2578 (30.63) | 13.50 (1.68) | 19205 (30.66) |
| Central | 10195 (18.80) | 11.47 (1.45) | 1341 (15.93) | 13.22 (1.63) | 11536 (18.41) |
| East | 5991 (11.05) | 12.29 (1.50) | 907 (10.78) | 14.04 (1.62) | 6898 (11.01) |
| Northeast | 5548 (10.23) | 11.71 (1.63) | 922 (10.96) | 13.68 (1.83) | 6470 (10.33) |
| West | 7967 (14.69) | 11.77 (1.57) | 1460 (17.35) | 13.56 (1.65) | 9427 (15.05) |
| South | 7904 (14.57) | 11.74 (1.53) | 1208 (14.35) | 13.33 (1.66) | 9112 (14.54) |
| Total | 54232 (86.57) | 11.72 (1.57) | 8416 (13.43) | 13.52 (1.69) | 62648 (100.00) |

**Table 2:** Summary measures of community-level factors (N=23,448)

| Characteristic | Mean (SD) | 25th percentile | 75th percentile |
| --- | --- | --- | --- |
| Proportion of women having completed formal education at least till 10th grade | 0.15 (0.15) | 0.04 | 0.22 |
| Proportion of households belonging to SC/ST group | 0.37 (0.32) | 0.10 | 0.57 |
| Proportion of households following Muslim religion | 0.11 (0.24) | 0.00 | 0.09 |
| Proportion of households in the poorest wealth category | 0.23 (0.27) | 0.00 | 0.38 |

In the bivariate Model 1 (Table 3), a one standard deviation (0·15) higher community-level women’s education was associated with (0·676(*β*)*0·15) 0·1 g/dl higher hemoglobin level, on average (p<0·001). Adding individual level covariates reduced the values of coefficients associated with the primary predictor (*β*=0·566 in model 2 and 0·481 in model 3), but retained the positive association of the primary predictor observed in Model 1 (p<0·001). Adding mother’s education (Model 4) further reduced the *β* (0·31), but retained the positive association between the primary predictor and the outcome (p<0·001).The average hemoglobin level in adolescents was higher by 0·035 g/dl if the proportion of women who were educated up to the tenth grade or above in a community was 15 percentage points higher (p<0·01) in the fully adjusted model (Model 5). Notably, nearly 45% of the PSUs in our sample had less than 10% of the women with a tenth-grade education. If 100% of the women in these communities acquire at least a tenth-grade education, our findings suggest that it might lead to (0·9*0·235(*β*)) 0·22 g/dl higher hemoglobin level on average, among all adolescents in those communities. Similarly, in nearly 71% of the PSUs having less than 20% of the women with a tenth-grade education, the gain in average hemoglobin level would be 0·19 g/dl for all adolescents in such communities. In all the models, unexplained variance at all the five levels remained statistically significant, indicating the importance of contextual factors at the household-, community-, district-, and state-levels.

**Table 3:** The association (regression coefficients and 95% CI) of community-level women’s education with adolescent hemoglobin level in unadjusted and adjusted models (N in all models = 62,648 adolescents)

| Characteristics | Model 1 | Model 2 | Model 3 | Model 4 | Model 5 |
| --- | --- | --- | --- | --- | --- |
| Fixed part | Coefficient (95% confidence interval) |  |  |  |  |
| Community-level women's education | 0.676 (0.561, 0.792)*** | 0.566 (0.460, 0.673)*** | 0.481(0.372, 0.590)*** | 0.310 (0.191, 0.427)*** | 0.235 (0.094, 0.377)** |
| Adolescent's age |  | 0.001 (0.000, 0.003)* | 0.001 (-0.001, 0.002) | 0.001 (-0.000, 0.002) | 0.001 (-0.000, 0.002) |
| Adolescent's female gender (Reference category: boys) |  | -1.792 (-1.828, -1.757)*** | -1.791(-1.826, -1.755)*** | -1.790 (-1.826, -1.755)*** | -1.790 (-1.826, -1.755)*** |
| Adolescent's education |  |  | 0.019 (0.014, 0.025)*** | 0.015 (0.010, 0.020)*** | 0.013 (0.008, 0.019)*** |
| Mother's education |  |  |  | 0.013 (0.009, 0.016)*** | 0.007 (0.003, 0.011)*** |
| Father's education |  |  |  |  | 0.003 (-0.001, 0.001) |
| Family size |  |  |  |  | -0.001 (-0.007, 0.004) |
| Social group<br>(reference category:<br>Scheduled Caste) |  |  |  |  |  |
| Scheduled Tribe |  |  |  |  | -0.103 (-0.155, -<br>0.050)*** |
| Other backward<br>classes |  |  |  |  | 0.088 (0.050,<br>0.126)*** |
| General |  |  |  |  | 0.081 (0.036,<br>0.126)*** |
| Religion (reference<br>category: Hindu) |  |  |  |  |  |
| Muslim |  |  |  |  | 0.059 (0.001,<br>0.117) |
| Christian |  |  |  |  | 0.127 (0.038,<br>0.216)*** |
| Other |  |  |  |  | -0.036 (-0.115,<br>0.042) |
| Household wealth<br>(reference category:<br>poorest) |  |  |  |  |  |
| Poorer |  |  |  |  | -0.006 (-0.046,<br>0.035) |
| Middle |  |  |  |  | 0.021 (-0.026,<br>0.068) |
| Richer |  |  |  |  | 0.011 (-0.043,<br>0.064) |
| Richest |  |  |  |  | 0.097 (0.033,<br>0.161)** |
| Proportion of SC and<br>ST households |  |  |  |  | -0.050 (-0.120,<br>0.020) |
| Proportion of Muslim<br>households |  |  |  |  | 0.074 (-0.014,<br>0.163) |
| Proportion of poorest<br>households |  |  |  |  | 0.016 (-0.076,<br>0.108) |
| Place of residence<br>(reference category:<br>Urban) |  |  |  |  |  |
| Rural |  |  |  |  | 0.025 (-0.016,<br>0.065) |
| Region (reference<br>category: North) |  |  |  |  |  |
| Central |  |  |  |  | -0.038 (-0.404,<br>0.327) |
| East |  |  |  |  | 0.769 (0.454,<br>1.085)*** |
| Northeast |  |  |  |  | 0.021 (-0.296,<br>0.338) |
| West |  |  |  |  | 0.195 (-0.158,<br>0.547) |
| South |  |  |  |  | 0.282 (-0.184,<br>0.748) |
| Constant | 11.905 (11.757,<br>12.052)*** | 13.189 (12.925,<br>13.452)*** | 13.200 (12.936,<br>13.464)*** | 13.148 (12.884,<br>13.411)*** | 12.919 (12.599,<br>13.238)*** |
| Random part | Variance (95% confidence interval) |  |  |  |  |
| State | 0.175 (0.100,<br>0.305) | 0.173 (0.100,<br>0.298) | 0.176 (0.103,<br>0.304) | 0.172 (0.100,<br>0.300) | 0.083 (0.045,<br>0.153) |
| District | 0.127 (0.110,<br>0.148) | 0.126 (0.110,<br>0.145) | 0.125 (0.108,<br>0.144) | 0.124 (0.108,<br>0.143) | 0.120 (0.103,<br>0.138) |
| Primary sampling unit | 0.198 (0.176,<br>0.222) | 0.146 (0.127,<br>0.166) | 0.145 (0.127,<br>0.166) | 0.145 (0.126,<br>0.166) | 0.141 (0.123,<br>0.162) |
| Household | 0.434 (0.375,<br>0.506) | 0.432 (0.377,<br>0.494) | 0.429 (0.375,<br>0.493) | 0.429 (0.373,<br>0.491) | 0.428 (0.373,<br>0.493) |
| Residual | 2.033 (1.971,<br>2.100) | 1.724 (1.667,<br>1.782) | 1.724 (1.667,<br>1.782) | 1.724 (1.667,<br>1.782) | 1.716 (1.665,<br>1.780) |

\*p<0.05, \*\*p<0.01, \*\*\*p<0.001

## Discussion

Our study of 62,648 adolescents aged 15 through 17 years found that community-level women’s education was positively associated with hemoglobin level in adolescents, accounting for relevant covariates and contextual levels. Further, we found evidence of the importance of context on adolescent hemoglobin. To our knowledge, this is the first attempt to examine the influence of contextual factors on adolescent health or nutritional status. These findings are especially relevant considering that 71% of communities in this nationally representative survey from India had only 20% or fewer women with a tenth-grade education.

Our findings are similar to the previous studies that examined the relationship of contextual determinants with child health outcomes in the Indian context. Kravdal’s 2004 study^12^ of 90000 Indian mothers of under-fives, from a nationally representative survey, demonstrated that the average years of education among women at the community-level was associated with lower child mortality (*β*-0·045, p<0·01) independent of mother’s education. Using 1994 data from 5,623 infants living in 195 districts of 16 states Parashar^13^ suggested that average female literacy rate at the district-level was associated with higher complete immunization status of children under-two independent of mother’s education (*β* 1·49, p<0·05). Burroway^14^ in 2016 showed that percentage of female secondary school enrolment at the country level reduced the odds of stunting in under-fives (*β* 0·991, p<0·05) across 50 developing countries. Although the size of the estimate for community-level women’s education was smaller in our study as compared to these previous studies, notably, its association with the outcome was in the expected direction. Furthermore, the coefficient’s estimate is not strictly comparable across the studies, as the operationalization of community-level women’s education, the outcome of interest, the study population and the nature of causal influence were different among all the studies. Moreover, adolescent health might be less influenced by community-level women’s education than the health of under-fives: adolescents might get affected by a higher number of influencers, so the change in the outcome influenced by one particular determinant might be smaller in comparison. However, as our findings are applicable to a large number of adolescents, even a small change in hemoglobin level of adolescents due to the influence of the context, has a greater effect at the population level.

Our findings are in line with recent evidence^15^ from rural Odisha, positing social and unequal gender norms in community that deprioritize women’s preventive health, especially in non-pregnant women of reproductive age, as important beyond individual-level factor, resulting in poor uptake of iron supplementation. Although the focus of their qualitative inquiry was on exploring barriers to iron supplementation in women,^15^ they have identified the need for addressing social norms, as extra-individual level factors in reducing anemia burden in India. Triangulating our quantitative evidence, derived from a nationally representative survey with qualitative understanding of barriers to anemia prevention,^15^ adds strength to our argument, that socio-cultural norms, valuing women’s well-being, as captured by community-level women’s education is an important determinant of adolescent hemoglobin.

Our investigation employed a measure of women in India completing education up to the tenth grade, which itself is a bar set too low. However, if this goal is met, our findings suggest that it will potentially lead to appreciable gains in the hemoglobin level of adolescents. Thus, ensuring the education of girls beyond the tenth grade might have an even greater influence on adolescent hemoglobin level. However, our core interest is in moving beyond increasing the number of years of completed schooling among women and directing attention to the socio-cultural norms in a community that values women and facilitates their completing high school (or more). We argue that community-level women’s education is a crude proxy for the strength of socio-cultural norms of a community that values women. Further, measuring it as the number of completed years of schooling fails to capture the quality of the education acquired, which is frequently of poor quality and inequitably distributed in India.^16^ Therefore, it is a weak proxy for capturing the health-promoting impacts of schooling, such as an enhanced critical thinking ability, empowerment conducive towards healthy behaviors, and a change in social norms. It is reported that curriculum in Indian schools lack in engaging children and instigating critical thinking in them.^17^ Despite the use of such a crude indicator, we were able to find modest statistical evidence supporting our hypothesis. Thus, despite the coefficient being small, it gives a valuable insight about the influence on adolescent hemoglobin level of community-level women’s education and all that this indicator represents.

Although we have not empirically examined the potential pathways through which this relationship operates, theoretically, such contextual impact^18^ is plausible as theorized in the case of child health, ^12,14,19^ and by extension might be relevant for adolescents. Education enhances women’s access to information, and widens her social network for instance through working outside home, exposing them to new knowledge and behavior norms such as those related to (adolescent’s) nutrition, ^13,14^ and education.^20^ Educated women are more likely to break from traditions in adopting advances in nutrition,^14^ and might be better equipped to use available resources to improve dietary quality ^21^ and nutritional status^5^ of their children. For instance, they might be more aware about dietary requirements and sources of nutrients including iron, and ensure its regular consumption among adolescent children.^22^ Educated women might be prompt in using health services for children and able to navigate health systems better.^14,19,22^ Education might enhance women’s participation in household decision making, increasing her bargaining power to divert resources towards nutrition, health and education of her children.^14^ Favorable attitudes and progressive behaviors towards adolescent health among a substantially higher number of educated women in the neighborhood likely exposes other community members to health-promoting choices made, and advocated, by educated women. Through processes such as social learning and social influence ^23^ it likely spreads to other community members including the adolescents and their mothers. Perhaps this alters social norms and expectations related to adolescent care, nutrition, health and education. Further, a critical number of educated women in the community may generate the social capital needed to change a community’s ability to better use the available health services and make local health workers more responsive to the demands of the community.^19^ Educated women might also contribute towards expansion of economic wealth creation in the community, leading to investments in health, sanitation and education.^12^ Thus altered normative expectations and community wealth creation by educated women do not simply benefit their offspring but could also benefit adolescents in the community.

While our focus was on the immediate neighborhood of residence as a community, (village/village cluster/urban wards) we also found evidence of the influence on adolescent hemoglobin of the context at state- and district-levels, even after accounting for a host of covariates. These findings underscore the potential relevance of context on adolescent health. This indicates that it is unrealistic to expect major improvement in adolescent health through attempts focused exclusively on attributes of adolescents without modifying the structural level determinants at higher levels.

Our findings suggest that policy interventions on adolescent health need to target contextual determinants of hemoglobin, and by extension anemia, in adolescents. India’s key intervention to reduce anemia burden-*Anemia Mukt Bharat* (AMB) noticeably prioritizes maximizing iron supplementation.^24^ Although essential, iron supplementation might be effective in reducing anemia to a limited extent, and in certain contexts.^25^ Notably, less than half of anemia in developing countries is attributable to iron deficiency.^26^ This might explain the ineffectiveness of India’s long history of iron supplementation in reducing anemia. Several other biological factors, such as poor bioavailability of iron, poor iron absorption, deficiency of other nutrients, exposure to recurrent infections and parasitic infestations, inadequate iron stores, and inadequate replacement of lost iron during menstruation in girls are involved in causing anemia.^27^ Drawing from the Ecosocial theory,^6^ these may be conceptualized as embodiments of poor living conditions and disadvantaged social positions. Addressing these issues call for structural and health behavior changes. Evidence suggests that investing in women’s education is among the best investments that could facilitate economic development, socio-political transformations, and improve the health of generations.^28^ We argue that such an investment is not just the means to an end (better nutrition of adolescents) but also an important outcome itself, which ideally reflects greater value placed on women’s well-being in general.

The AMB program also includes intensive behavior change communication^24^, however, it is targeted at individual-level. Adolescence is a transitional phase in which increasing autonomy and time spent outside the home, increase potential influence of peers and others outside the family on adolescent’s behaviors.^29^ While creating awareness related to health and nutrition in adolescents is crucial, we argue that an individual-level approach is insufficient to realize effective behavior change. Poor uptake of iron supplementation in urban girls despite efforts to increase awareness,^30^ and motivation from parents, teachers and friends increasing its uptake in girls on the other hand^31^ suggests the same. We need to consider adolescents as social beings embedded in their communities^6^, who need support from the community for engaging in health enhancing behaviors. Behavior change efforts need to emphasize modifying the local context within which adolescents make decisions. Our findings suggest that raising the level of women’s education could be an important factor facilitating this process.

There is evident gender gap in literacy rates and enrolment ratios at every level of formal education in India; and is more pronounced among socio-economically marginalized women.^32^ Our findings advocate for the strengthening of policies encouraging women’s education, with a special focus on socio-economically disadvantaged women, at every level of formal education. In India, nearly 40% of girls aged 15 to 18 years are out of school.^33^ Discriminatory gender norms valuing and prioritizing boy’s education over girl’s education, poverty, early marriage and child bearing, viewing girls as liabilities, dowry practices, child labor, household work, and care responsibilities are important demand-side barriers to girls’ education.^17^ Whereas evaluation of the supply side points towards an insufficiently funded education system, with severe shortage of secondary schools and teachers especially in rural areas.^33^ Not having a school nearby increases the opportunity cost of sending girls to school, which puts poor girls at double disadvantage: because of gender and poverty. Concerns over girls’ safety and security is also an important barrier in girls’ education.^34^ Thus, women’s education is a consequence of structural level factors, social norms and attitude towards women’s education, parental attitudes and household level factors that collectively determine availability and access to education for women. This therefore points towards an urgent need for a convergent action, directed towards structural and socio-cultural transformations that support and facilitate women’s education beyond elementary level.

Our study has several limitations. We included a narrow age-range of adolescents in our analysis because of the data constraints. It is suggested that early (10 to 14) and late (15-19) adolescence differ in health status and its determinants. It is possible that younger adolescents (10 to 14 years), who likely have less autonomy than the older adolescents, could be more affected by several aspects related to norms of care, feeding practices, and other health behaviors, in the family; which in turn are arguably affected by the broader socio-cultural norms and behaviors of the educated women in the community. We therefore submit that our findings represent a conservative estimate of the impact of community-level women’s education on adolescent health. Further, because of the cross-sectional nature of the data, the association does not imply any directionality or causal relationships. If the association is presumed to be causal, why and how remains unaddressed. Relative importance of different processes and pathways that we theorized, as well as the role of other contextual determinants remain to be empirically tested, all of which could indicate the most rewarding directions to focus our efforts. Next, our analytical sample mainly consisted of unmarried girls (85%), living with both the parents in the same household. Only 15% of the total households selected for the survey were eligible for measuring hemoglobin level in adolescent boys as per the survey design.^8^ This limited the representation of boys in our sample. As we aimed to adjust for parental education, we excluded married adolescents and those adolescents whose father did not stay in the same household as information on parent’s education was not available for these adolescents. Our findings therefore are not generalizable to married adolescents or who are not staying with both their parents. We however theorize that community-level women’s education might have a greater influence on the health of this subgroup of adolescents, than what we found in our sample. Lastly, while large-scale national-level studies could hint towards a broader picture and a general pattern, given that local contextual level factors affect their health, future research needs to emphasize gaining deeper insight into how adolescents as a diverse group have different health needs. This requires a detailed enquiry based on a sound theoretical premise.

The strength and novelty of our findings lie in demonstrating the contextual influences at various population levels on adolescent hemoglobin while providing policy-relevant evidence of the association of a contextual factor such as community-level women’s education with adolescent hemoglobin in the Indian context.

We conclude by reiterating that although the role of social determinants at multiple contextual levels in adolescent health and nutrition is emphasized in literature,^29^ policy action aiming at adolescent health in India is directed at individual-level biological determinants. Our findings bring the role of contextual factors to the center, highlighting the importance of structural changes and the need to engage communities along with adolescents. This might serve to fill a crucial link in our efforts to improve adolescent health and nutrition.

## Data Availability

We have used publicly available, National Family Health Survey India 2015-16 data: International Institute for Population Sciences (IIPS) and ICF. National Family Health Survey (NFHS-4) 2015-16 [Dataset] Accessed from https://dhsprogram.com/ on 7/11/2019. For query contact Ankita R. Shah at

## Acknowledgements

ARS thanks Anupam Sharma and Krunal Shah for their help in putting together analytical dataset.

